# Confirmation of male well-being indicators in Malaysia

**DOI:** 10.1101/2023.11.05.23298123

**Authors:** Siti Zaiton Mohd Ajis, Emma Mohamad, Arina Anis Azlan

**Affiliations:** Centre for Research in Media and Communication, Faculty of Social Sciences and Humanities, Universiti Kebangsaan Malaysia, Bangi, Malaysia; UKM x UNICEF Communication for Development Centre in Health, Faculty of Social Sciences and Humanities, Universiti Kebangsaan Malaysia, Bangi, Malaysia

## Abstract

**Background:** Assessing well-being can be tricky due to its subjective nature which may result in inaccurate or incomplete evaluations. This is particularly challenging in measuring male well-being, as traditional gender roles and expectations often discourage normalising discussions about men’s health concerns. Studies reveal notable obstacles in the way men perceive, behave, and hold beliefs about their health and well-being which may result in underreporting of health issues among men. A gender-specific measurement of well-being for men is therefore essential and merits further examination.

**Methods:** This study aims to validate a male well-being instrument in the context of Malaysian men using confirmatory factor analysis (CFA). An online cross-sectional survey involving a total of 651 Malaysian men aged 18 and above was conducted utilising a 33-item male well-being instrument developed in a preliminary study.

**Results:** The analysis resulted in a satisfactory 24-item model with six dimensions: self-confidence (4 items); family/close relationships adaptation (5 items); physical health (3 items); living environment adaptation (4 items); autonomy and agency (4 items); and economic stability (4 items). There were high correlations among the 24 items. The internal consistency reliability was robust, with no floor or ceiling effects. These results represented equivalence and consistency among the responses to items, suggesting that the items were homogenous in measuring Malaysian male well-being.

**Conclusions:** This study confirms the suitability of a 24-item instrument measuring male well-being in Malaysia. The instrument may possibly be used in similar Asian cultures as it achieved strong reliability, structural validity and construct validity that fulfilled goodness-of-fit criteria.

## Introduction

Well-being is an expression that is often used to describe what is good or bad for an individual’s life. Aspects of well-being such as a being comfortable, content, and happy have subjective definitions. This makes the concept of well-being difficult to define, encompassing various aspects with various techniques in its measurement. Until now, the measurement of the concept of well-being is still a matter of discussion because it involves many dimensions of life that can be measured from various angles [1].

The measurement of well-being is an important subject to discuss because a country uses the measurement of well-being to measure the effectiveness of the implementation of a country’s social and economic development policies whether on individuals, families, or communities. Measuring well-being can also be used specifically as a guide for government programs to improve the quality of life among specific groups [2].

The subjective and objective measurement of well-being has been widely discussed in forming well-being indexes abroad and in Malaysia. There are also debates surrounding objective and subjective measures of well-being because the differences between the two are distinct [3–9].

The concept of quality of life is considered as an objective measurement, while well-being is aimed at subjective measurement of quality of life and is better known as subjective well-being [10]. The relationship between the two is also sometimes seen from various points of view, for example, there are parties who accept and use them interchangeably [11]. There are even researchers who use the term life satisfaction to describe the state of quality of life and subjective well-being [12,13].

Self-assessment is often used to gauge the level of individual well-being. However, with the use of self-assessment measurement methods, questions arise about the exact method of determining well-being such as whether self-assessment or objective assessment from a third party is more appropriate to use. In addition to that, the assessment of perception by different genders gives different perceived values of well-being [14].

The assessment of the perception of well-being also varies according to culture [15]. Therefore, the formation of standards or well-being benchmarks need to take into account gender and cultural norms of the group being studied. This study focuses on well-being in the context of men because it recognizes that gender and sociocultural influences that emphasize aspects of masculinity affect men’s behaviour in daily life and understanding of their environment.

In addition, norms that shape men’s responsibilities affect men’s perceptions of masculinity, the value they place on well-being and their behaviour in seeking that well-being [16]. In the traditional norms setting, men are considered leaders in family as well as in the social community and the country. Studies have found that conventional Malay men express their feelings of love through their commitment as academic supervisors, personal advisors, financial contributors, educators, positive role models, maintainers of discipline, and spiritual leaders [17,18]. From an Islamic perspective, studies have shown that culturally-bound veterans in Malaysia come from a collectivistic culture, where Islam as their religion and the way of life affects the Malay culture. As Islam is embedded in a Muslim’s way of life and beliefs, it also permeates their values, behaviour, and way of thinking [19,20]. This matter is often debated when it comes to measuring well-being because of the existence of gender differences and norms in individuals, society, and even the country itself. Therefore, understanding the influence of social norms on communication and behaviour is crucial for promoting positive health outcomes and supporting men in their unique needs and preferences.

This study focuses on well-being in the context of men because it acknowledges that gender and sociocultural influences emphasizing aspects of masculinity affect men’s behaviour in daily life and understanding of their environment. The aim of the study is to address this gap by developing a comprehensive instrument tailored to the context of well-being among men in Malaysia. By focusing on specific dimensions and indicators that are culturally and contextually appropriate, the instrument seeks to provide a nuanced understanding of personal well-being experiences among Malaysian men. Thus, in the context of this study, the researchers use a self-rated, 33-item male well-being instrument proposed by Ajis et al. [21] to observe how men subjectively view their lives and propose a set of items suitable to measure male well-being in the Malaysian context. Consequently, well-being dimensions and indicators appropriate for use among Malaysian men were identified and tested.

## Materials and method

### Study design

An earlier population-based cross-sectional study [21] has assessed the suitability of male well-being indicators for the Malaysian context. The researchers developed and tested a 33-item male well-being scale in Bahasa Melayu and English. Adapted measures from this instrument were utilized in the present study. A two-level face validation process was conducted to validate the 33 items measuring Malaysian male well-being. Some items were also reworded upon recommendation by health communication experts through the face validation stage to allow for better comprehension and reduce confusion for respondents. A cross-sectional survey was conducted among the Malaysian male population to validate the male well-being indicators and ensure that the instrument reflects the country’s male citizens. Participants were aged 18 and above and resided in Malaysia.

### Ethical approval

This study was submitted for ethical review and received approval from the Research Ethics Committee from the National University of Malaysia (UKM) which governs all medical/health/science/social-related research in UKM. The ethical approval number is UKM PPI/111/8/JEP-2020-43.

All respondents were above 18 years old and therefore the study involved no minors. All respondents also agreed with the online written consent form that clearly stated their rights and the nature of participation in the study before being asked to answer the survey. This online consent form was also submitted and approved by the Research Ethics Committee, UKM.

### Sampling method

The present study involved the Malaysian male population aged 18 and above. Data collection was conducted online during the COVID-19 period which limited the study’s sampling technique. In order to obtain respondents, the study utilised convenience and snowball sampling techniques based on several inclusion criteria (i.e., male, Malaysian, aged 18 and above, residing in Malaysia) and used professional and personal networks to reach as many male respondents as possible.

The sample size was calculated based on the number of items formed and the total male population in Malaysia. By using sample size calculation based on total items by Chua [22] and sample size determination by Krejcie and Morgan [23] and Bukhari [24], a minimum sample size of N=549 respondents was required in this study. In addition, the sample calculation also considers an 80 percent response rate, so at least 659 questionnaires were distributed to obtain a minimum of 549 respondents. This sample size is sufficient to perform confirmatory factor analysis (CFA) with a 95% confidence level on the model formed and is sufficient to represent the Malaysian male population.

Data collection was conducted for four months between 1st January 2021 to 30th April 2021 using the Survey Monkey platform. Respondents took an average of 10–15 minutes to fill in the questionnaire.

### Instrument

The Ajis et al. [21] male well-being instrument was adapted to obtain respondent assessment of personal well-being. The questionnaire contained 33 items measuring personal well-being on a seven-point bipolar scale (1=strongly disagree, 7=strongly agree). Respondents answered the questions by indicating their level of agreement with each statement. The 33-item male well-being model was formed through exploratory factor analysis and obtained a very good level of internal consistency where the Cronbach’s alpha of all well-being dimensions were at values above .70 [21], meeting the level of reliability suggested by Bond and Fox (25). The dimensions and items that make up the construct of personal well-being (Fig 1) are self-confidence (8 items); family/close relationships adaptation (8 items); physical health (3 items); living environment adaptation (5 items); autonomy and agency (4 items); and economic stability (5 items) and will be referred to hereafter as the original measurement model in this study.

**Fig 1.**
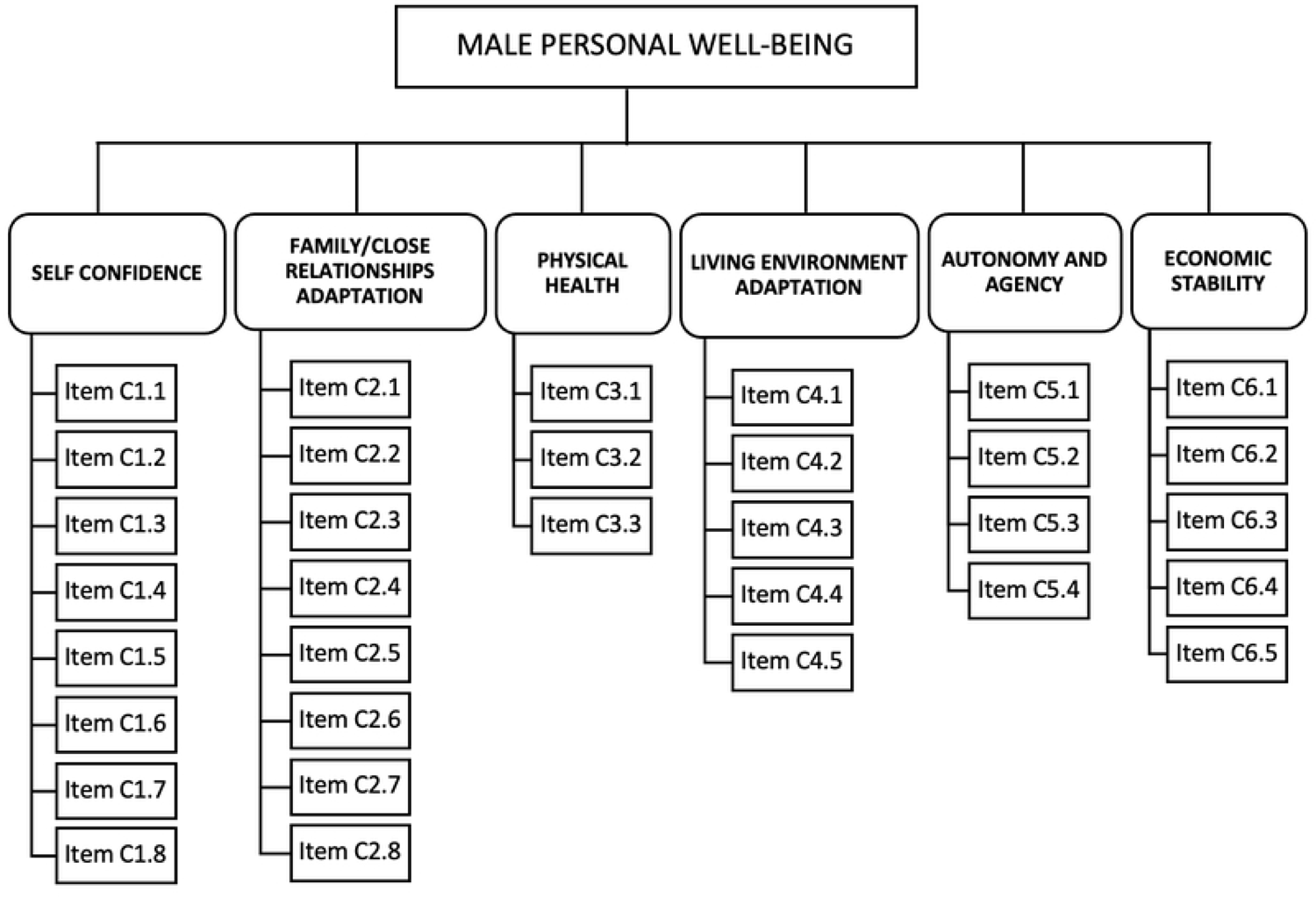
Original Measurement Model hypothesized (Ajis et al. 2021).

### Participant recruitment and data collection procedure

Professional and personal networks were used to distribute the self-reported questionnaire to respondents during the COVID-19 pandemic. The two main platforms used in disseminating these online survey links were social media (Facebook, Twitter, and Instagram) and messaging platforms (WhatsApp). A general overview of the questionnaire was first given in a WhatsApp/social media message post followed by online survey links to Malay and English versions of the questionnaire. A digital consent form was included in the survey and each participant needed to click the agreement button as a sign of consent to participate in the survey. Although the researchers aimed to collect 659 responses, a total of 851 respondents participated in the online questionnaire throughout the data collection period. However, the data cleaning process found that 200 respondents did not meet the study criteria and were removed (female [n=143], non-Malaysian citizen [n=43], answered all questions with the same answer [n=12] and extreme outliers in the normality test [n=2]). A total of 651 complete responses with no missing data were obtained and analyzed.

### Data analysis

The data collected in this study were analyzed using the Statistical Package for the Social Sciences (SPSS) software and AMOS version 26.0. In this study, the research data was normally distributed with all variables obtaining skewness and kurtosis values between −1.107 to 1.507. In the context of this study, the items to be tested are self-assessment items on the dimensions of male well-being construct. Confirmatory factor analysis (CFA) was conducted for all dimensions to validate instruments measuring the male well-being construct in terms of unidimensionality, validity, and reliability [26–28]. The original measurement model must meet three types of validity: convergent validity, construct validity, and discriminant validity [26–31]. The fit of the data to the model was examined using goodness-of-fit indices, including (i) Absolute fit: root mean square error of approximation (RMSEA) and goodness-of-fit index (GFI); (ii) Incremental fit: adjusted goodness-of-fit index (AGFI), comparative fit index (CFI), Tucker–Lewis index (TLI), and normed fit index (NFI); (iii) Parsimonious fit; Chi-Square/Degree of freedom (Chisq/df). To assess reliability, the composite reliability of the construct was examined. Internal consistency was tested with Cronbach’s alpha, and values greater than or equal to 0.7 indicate satisfactory reliability.

## Results and Discussion

The purpose of this study was to test and confirm the Ajis et al. [21] 33-item male well-being instrument and propose a set of items to measure male well-being in the Malaysian context. The 33-item male well-being instrument was designed to measure the multiple aspects of male well-being in the Malaysian context, represented by self-assessment of well-being dimensions, namely self-confidence, physical health, autonomy and agency, economic stability, family/close relationship adaptation, and living environment adaptation. A total of 651 complete responses with no missing data were obtained and analyzed. Structural equation modeling was used to conduct a confirmatory factor analysis (CFA) on the dimensions of male well-being and define its structure.

### Participant characteristics

Table 1 shows that the study respondents consisted of various demographic backgrounds. Out of the total 651 male respondents, the average age was 33 years, indicating that most of the respondents were males from the Generation Y (27-44) group, representing half of the sample at 348 respondents (53.5%). The majority of the respondents were Malay (76.0%), followed by respondents who were Chinese (13.4%), and Indian (5.2%). The majority of respondents involved in this study were married (56.2%), had an undergraduate degree (35.8%), and had an estimated family income below RM4360 (55.9%).

**Table 1.** Characteristics of participants Variables.

| Variables | *n* | % |
| --- | --- | --- |
| Age | Mean = 33.87 (24) |  |
| Generation Z (18-26) | 193 | 29.6 |
| Generation Y (27 – 44) | 348 | 53.5 |
| Generation X (45-55) | 95 | 14.6 |
| Baby Boomers (56 – 71) | 15 | 2.3 |
| Ethnic Group |  |  |
| Malay | 495 | 76.0 |
| Chinese | 87 | 13.4 |
| Indian | 34 | 5.2 |
| Bumiputera Sabah/Sarawak | 33 | 5.1 |
| Serani | 1 | 2 |
| Bugis | 1 | 2 |
| Highest Education Level |  |  |
| UPSR / Equivalent | 8 | 1.3 |
| SRP / PMR / PT3 / Equivalent | 6 | 0.9 |
| SPM / SPMV / Equivalent | 98 | 15.1 |
| STPM / Diploma / Equivalent | 195 | 30.0 |
| Skill Certificate | 38 | 5.8 |
| Bachelor Degree | 233 | 35.8 |
| Master Degree/PhD | 73 | 11.2 |
| Marital Status |  |  |
| Single | 270 | 41.5 |
| Married | 366 | 56.2 |
| Divorced | 13 | 2.0 |
| Widowed | 2 | 0.3 |
| Estimated Household Income |  |  |
| RM 4360 below | 364 | 55.9 |
| RM 4,361 – RM 9,619 | 175 | 26.9 |
| RM 9,621 above | 112 | 17.2 |

### Construct validity of the male well-being instrument

Confirmatory factor analysis (CFA) was used to determine the fit of the original hypothesized model and check the reliability and validity of the measurement items. IBM SPSS AMOS version 26.0 was used for the procedure analysis of model quality and fit. Fig 2 shows the CFA output of the original measurement model hypothesized. Based on the output in Fig 2, the study needs to assess the three types of validity: construct validity, convergent validity, and discriminant validity together with composite reliability for male well-being construct. The construct must achieve all validity and reliability requirements before it can be released into practice.

**Fig 2.**
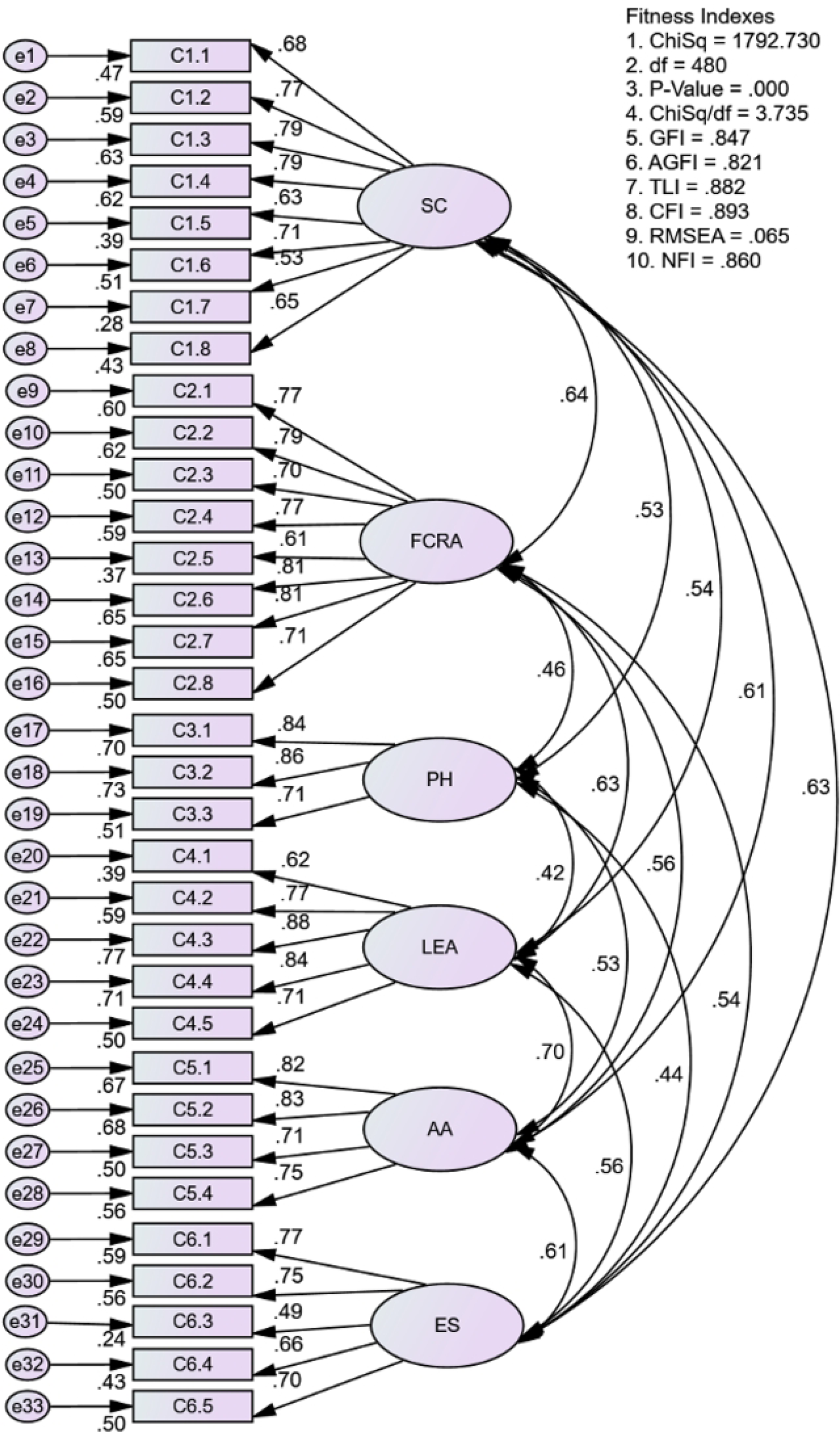
The original measurement model for male well-being construct.

The construct validity is achieved when the model achieves all three types of model fit categories: Absolute Fit (RMSEA < 0.08; GFI > 0.90), Incremental Fit (AGFI, CFI, TLI, NFI > 0.90) and Parsimonious Fit (ChiSq/df < 3.0) (26–31).

The assessment for construct validity is presented in Table 2. Based on Table 4, the test results indicate that the model fit does not fully adhere to goodness-of-fit indices. The analysis resulted in absolute fit; RMSEA = 0.065, GFI = 0.847, incremental fit; AGFI = 0.821, CFI = 0.893, TLI = 0.882, NFI = 0.860, and parsimonious fit χ2/df = 3.735 showing that the required levels are not fully achieved. Thus, the study concludes that the convergent validity of the original measurement model has not been achieved.

**Table 2.** Construct validity of original measurement model.

| Name of category | Name of index | Index value | Comments |
| --- | --- | --- | --- |
| 1. Absolute Fit | RMSEA | 0.065 | The required level is achieved |
|  | GFI | 0.847 | The required level not achieved |
|  | AGFI | 0.821 | The required level not achieved |
| 2. Incremental Fit | CFI | 0.893 | The required level not achieved |
|  | TLI | 0.882 | The required level not achieved |
|  | NFI | 0.86 | The required level not achieved |
| 3. Parsimonious Fit | Chisq/df | 3.375 | The required level not achieved |

RMSEA: Root Mean Square of Error Approximation
GFI: Goodness of Fit Index
AGFI: Adjusted Goodness of Fit
CFI: Comparative Fit Index
TLI: Tucker-Lewis Index
NFI: Normed Fit Index
Chisq/df: Chi Square/Degrees of Freedom

Next, the assessment for the convergent validity and composite reliability are shown in Table 3, while the discriminant validity among dimensions is shown in Table 4. The model had validity concerns due to AVE values for two dimensions (economic stability and self-confidence) not exceeding the threshold value of 0.5 which indicates the convergent validity for the original measurement model has not been achieved. However, the values of CR in Table 3 exceeded 0.6 which indicates that composite reliability for the original measurement model has been achieved [26–31]. The values for discriminant validity in Table 4 indicate that MSV and ASV values are smaller than AVE value and this indicates good discriminant validity for the original measurement model.

**Table 3.** Convergent validity and composite reliability of original measurement model for male well-being.

| Dimensions | CR | AVE |
| --- | --- | --- |
| Economic stability | 0.809 | 0.464 <sup>1</sup> |
| Family and close relationships adaptation | 0.91 | 0.56 |
| Self-confidence | 0.883 | 0.488 <sup>1</sup> |
| Physical health | 0.846 | 0.649 |
| Living environment adaptation | 0.876 | 0.589 |
| Autonomy and agency | 0.858 | 0.603 |

<sup>1</sup>: AVE less than 0.50.
CR: Composite reliability
AVE: Average variance extracted

**Table 4.** Discriminant validity among dimensions of original measurement model for male well-being.

| Dimensions | AVE | MSV | ASV |
| --- | --- | --- | --- |
| Economic stability | 0.464 <sup>1</sup> | 0.399 | 0.314 |
| Family and close relationships adaptation | 0.56 | 0.41 | 0.325 |
| Self-confidence | 0.488 <sup>1</sup> | 0.41 | 0.351 |
| Physical health | 0.649 | 0.28 | 0.23 |
| Living environment adaptation | 0.589 | 0.483 | 0.332 |
| Autonomy and agency | 0.603 | 0.483 | 0.364 |

<sup>1</sup>: AVE less than 0.50.
AVE: Average variance extracted
MSV: Maximum shared variance
ASV: Average shared variance

Fig 3 presents the measure of correlation among the six dimensions measuring male well-being. The analysis found none of the correlation values between any two dimensions, as indicated by double-headed arrows, exceeded 0.85. Thus, the model does not have multicollinearity problems.

**Fig 3.**
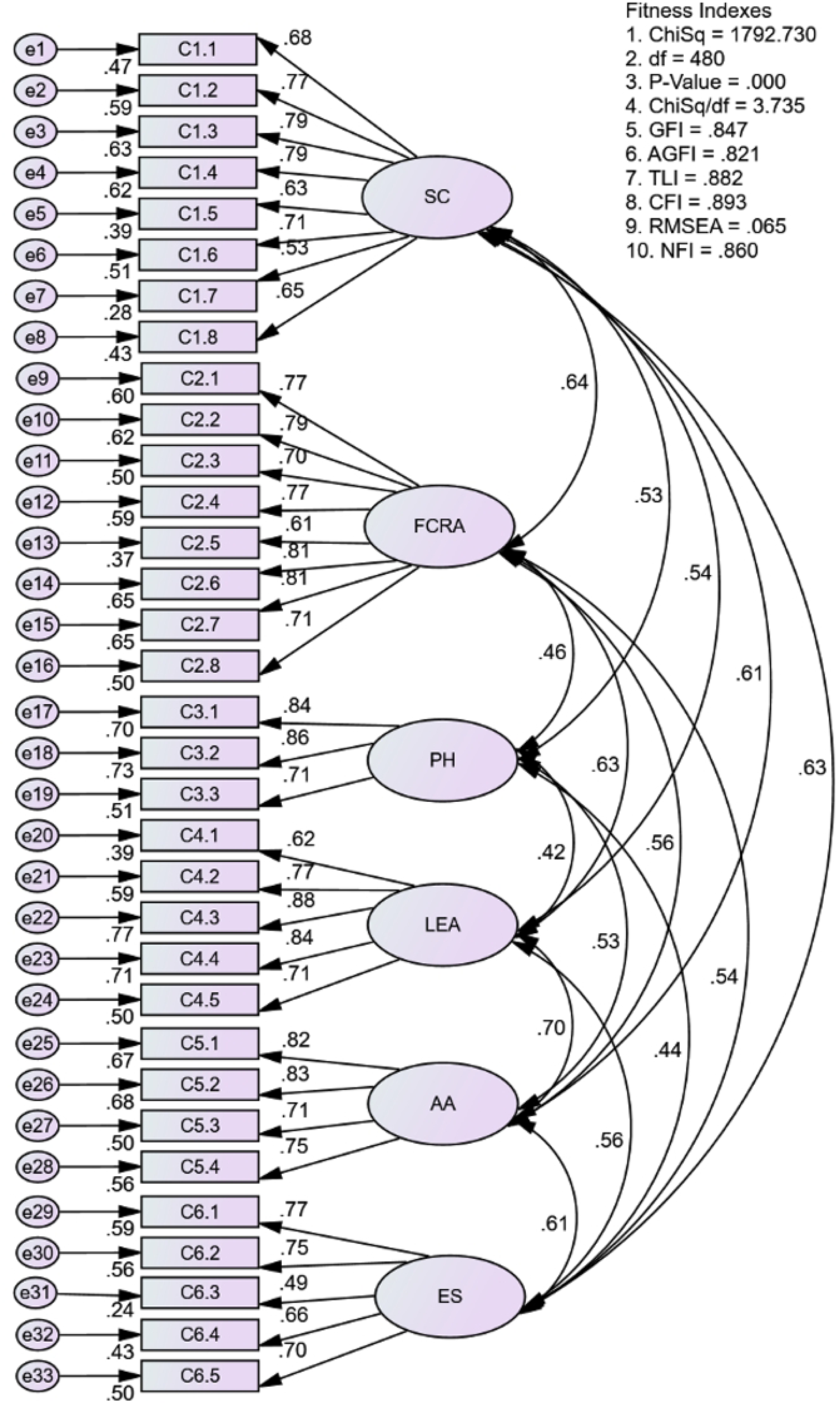
Correlation between dimensions measuring male well-being.

The correlation among dimensions obtained from Fig 3 are tabulated in Table 5. The diagonal values are the square root of the respective AVE while other values are the correlation between any two dimensions. Since all diagonal values are greater than any other values in the rows and columns, it can be concluded that the discriminant validity for the construct has been achieved [26–31].

**Table 5.** Discriminant validity index summary for the original measurement model.

| Dimensions | ES | FCRA | SC | PH | LEA | AA |
| --- | --- | --- | --- | --- | --- | --- |
| ES | 0.681 |  |  |  |  |  |
| FCRA | 0.539 | 0.748 |  |  |  |  |
| SC | 0.632 | 0.640 | 0.699 |  |  |  |
| PH | 0.445 | 0.465 | 0.529 | 0.805 |  |  |
| LEA | 0.558 | 0.629 | 0.539 | 0.424 | 0.768 |  |
| AA | 0.610 | 0.559 | 0.613 | 0.525 | 0.695 | 0.776 |

EC: Economic Stability
FCRA: Family and close relationships adaptation
SC: Self-confidence
PH: Physical health
LEA: Living environment adaptation
AA: Autonomy and agency

Even though the original measurement model indicates good discriminant validity and achieved composite reliability, the model fit needs to be improved to comply with the goodness of fit indices and display good construct and convergent validity. Since model fitness indexes did not meet the requirement level, the researchers examined the factor loadings for item removal. As shown in Fig 3, the factor loadings for item C1.7 (from dimensions self-confidence) and item C6.3 (from dimensions economic stability) were below the minimum value of 0.6 [26–28] and were therefore removed. A CFA was run for the second time with these items excluded. Fig 4 shows the new CFA findings. Only fitness indexes for RMSEA achieved the required level, even though the factor loading values for all items exceeded 0.6.

**Fig 4.**
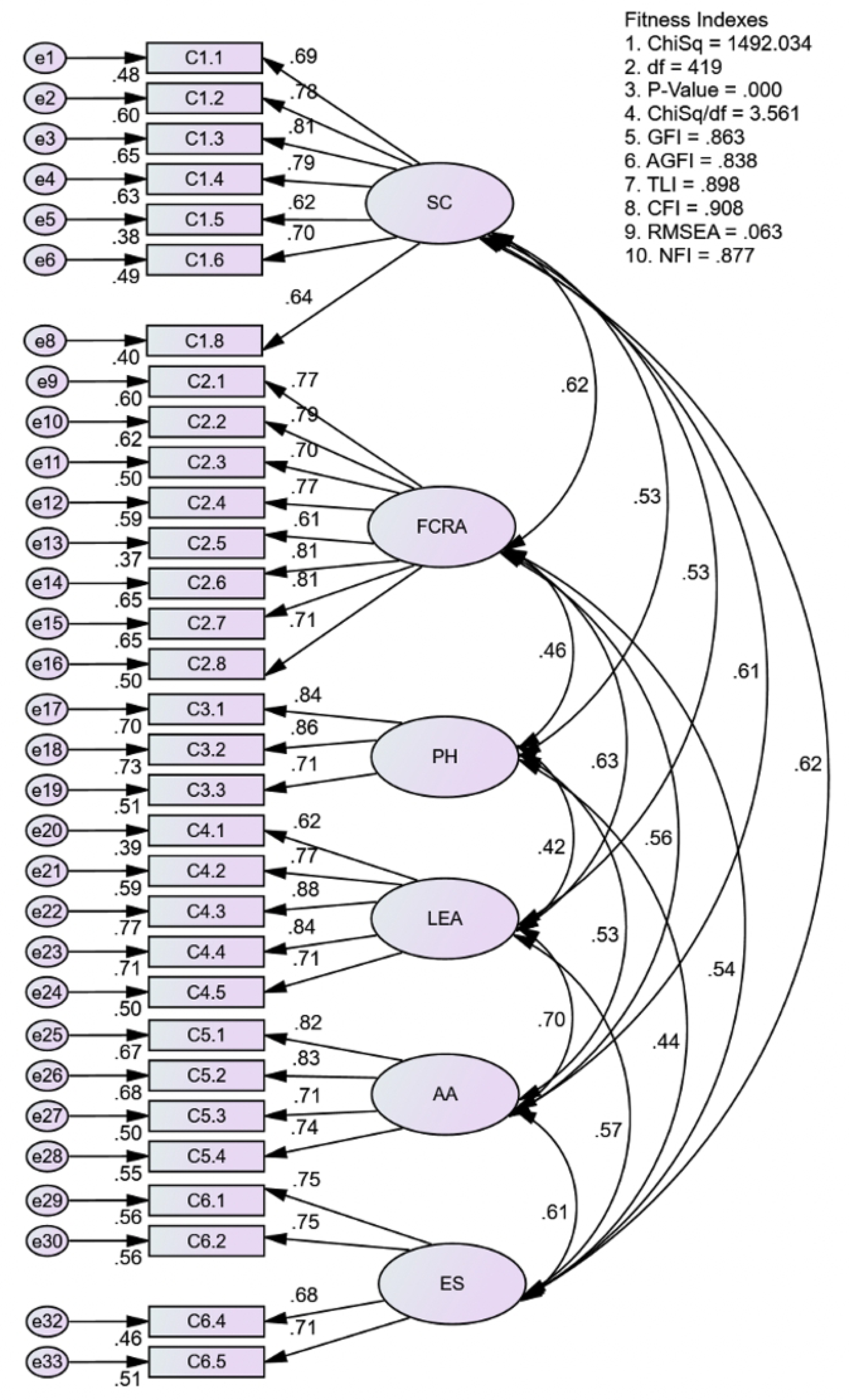
Factor loading and new fitness indexes after two items removed.

The model fitness indexes still had not met the required levels after selected items were removed. Therefore, the researchers identified redundant pairs of items through the modification index (MI). Table 6 indicates the highest covariance value, MI = 58.277 occurs between the errors e13 and e11; M= 57.27 occurs between the errors e28 and e27; M = 41.95 occurs between the errors e21 and e20; M = 40.766 occurs between the errors e10 and e9; M = 30.174 occurs between the errors e16 and e14; M = 28.423 occurs between the errors e8 and e3; M = 22.688 occurs between the errors e6 and FCRA; and M = 15.836 occurs between the errors e5 and FCRA. Based on the high covariance values in Table 6, the researchers constructed eight of modification models individually until the modification model complied with the goodness of fit indices.

**Table 6.** List of modifications made to the original measurement model.

| Items |  |  | MI | Par Change | Comment |
| --- | --- | --- | --- | --- | --- |
| e5 | <--> | FCRA | 15.836 | 0.079 | **8th Modification:** Delete item C1.5<br>MI > 15 shows item C1.5 and FCRAadapt are redundant |
| e6 | <--> | FCRA | 22.688 | 0.092 | **7th Modification:** Delete item C1.6<br>MI > 15 shows item C1.6 and FCRAadapt are redundant |
| e6 | <--> | e3 | 17.108 | -0.089 |  |
| e6 | <--> | e5 | 20.253 | 0.116 |  |
| e8 | <--> | FCRA | 15.021 | 0.082 |  |
| e8 | <--> | e3 | 28.423 | -0.126 | **6th Modification:** Delete item C1.8<br>MI > 15 shows item C1.8 and C1.3 are redundant |
| e8 | <--> | e6 | 19.787 | 0.123 |  |
| e21 | <--> | e20 | 41.95 | 0.18 | **3rd Modification:** Delete item C4.1 due to many redundant<br>MI > 15 shows item C4.2 and C4.1 are redundant |
| e22 | <--> | e20 | 20.018 | -0.109 | MI > 15 shows item C4.3 and C4.1 are redundant |
| e24 | <--> | e20 | 20.65 | -0.163 | MI > 15 shows item C4.5 and C4.1 are redundant |
| e28 | <--> | e27 | 57.27 | 0.234 | **2nd modification:** Constrain items C5.4 and C5.3<br>MI > 15 shows item C5.4 and C5.3 are redundant |
| e29 | <--> | e20 | 25.712 | 0.19 | MI > 15 shows item C6.1 and C4.1 are redundant |
| e32 | <--> | e20 | 17.306 | -0.152 | MI > 15 shows item C6.4 and C4.1 are redundant |
| e9 | <--> | LEA | 16.683 | 0.063 | MI > 15 shows item C2.1 and LEAdapt are redundant |
| e9 | <--> | AA | 17.949 | -0.088 | MI > 15 shows item C2.1 and AutAgency are redundant |
| e9 | <--> | e21 | 18.236 | 0.085 | MI > 15 shows item C2.1 and C4.2 are redundant |
| e9 | <--> | e33 | 23.347 | 0.161 | MI > 15 shows item C2.1 and C6.5 are redundant |
| e10 | <--> | e9 | 40.766 | 0.113 | **4th modification:** Delete item C2.1 due to many redundant<br>MI > 15 shows item C2.5 and C2.1 are redundant |
| e13 | <--> | e24 | 17.517 | 0.192 |  |
| e13 | <--> | e9 | 17.692 | -0.142 |  |
| e13 | <--> | e11 | 58.277 | 0.359 | **1st Modification:** Delete item C2.5.<br>MI > 15 shows item C2.5 and C2.3 are redundant |
| e16 | <--> | AA | 22.579 | <u>0.117</u> |  |
| e16 | <--> | e14 | 30.174 | 0.123 | **5th Modification:** Delete item C2.8<br>MI > 15 shows item C2.8 and C2.6 are redundant |

e: error indicator
MI: Modification Indexes
FCRA: Family and close relationships adaptation
LEA: Living environment adaptation
AA: Autonomy and agency

As a result, the researchers discarded seven items (C1.5, C1.6, C1.8, C2.1, C2.5, C2.8, and C4.1) and constrained items C5.4 and C5.3. Fig 5 shows the new CFA findings of the final modification model. Based on Fig 5 and Table 7, the test results indicate that the model fit fully adheres to goodness-of-fit indices. The analysis resulted in absolute fit; RMSEA = 0.049, GFI = 0.928, incremental fit; AGFI = 0.909, CFI = 0.957, TLI = 0.950, NFI = 0.932, and parsimonious fit χ2/df = 2.588 indicating achievement of required levels. Thus, it is concluded that the convergent validity of the modification model has been achieved.

**Fig 5.**
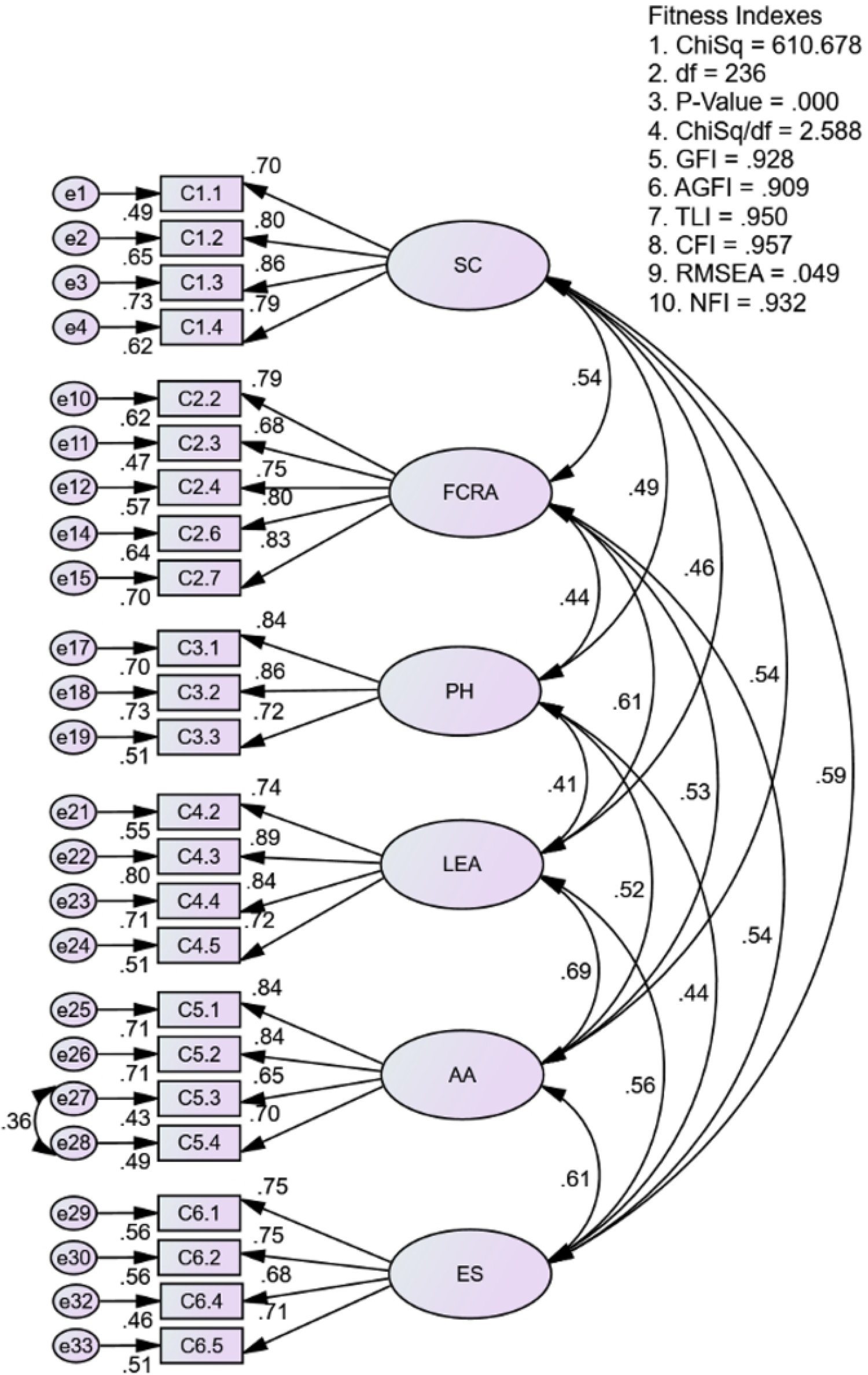
The final modification model for the construct of male well-being.

**Table 7.**
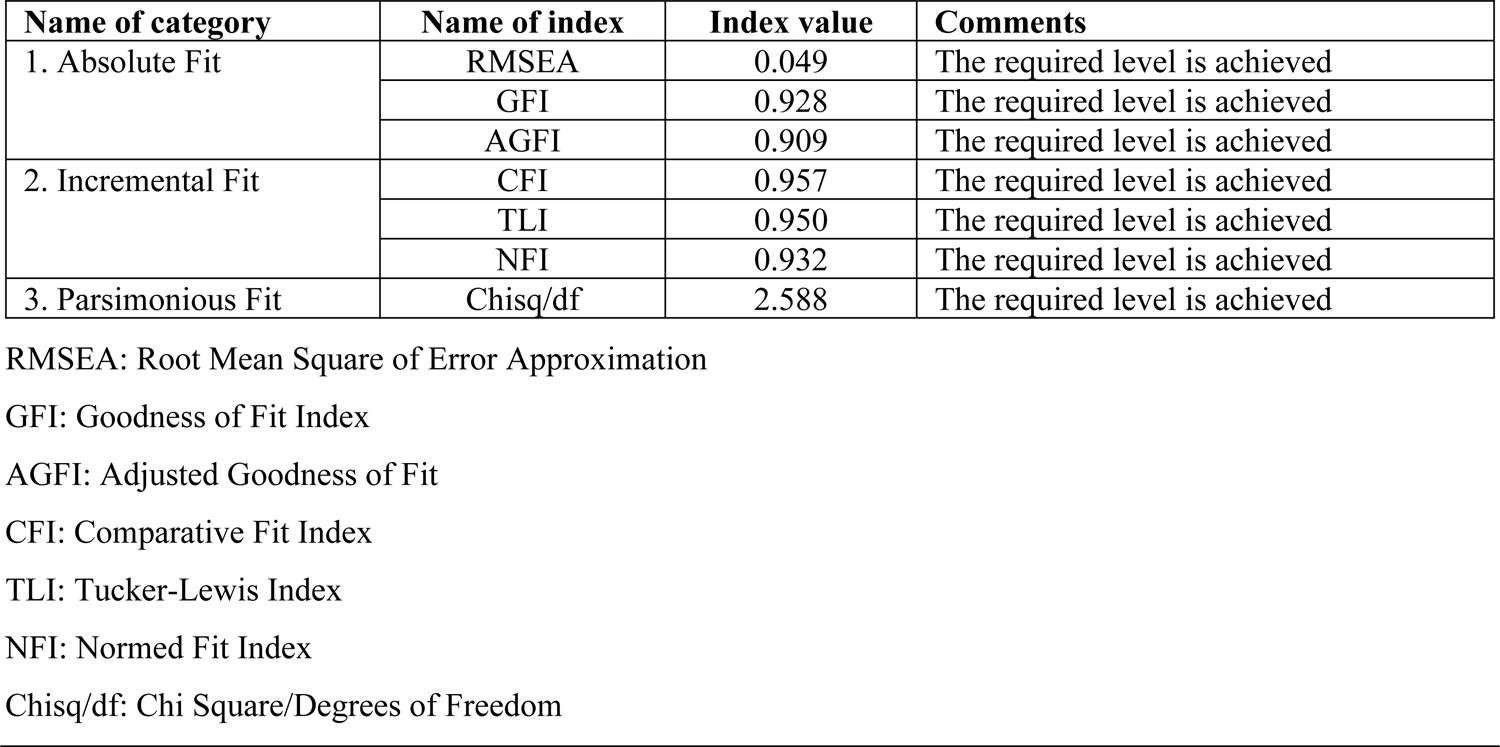
Construct validity of the final modification model.

| Name of category | Name of index | Index value | Comments |
| --- | --- | --- | --- |
| 1. Absolute Fit | RMSEA | 0.049 | The required level is achieved |
|  | GFI | 0.928 | The required level is achieved |
|  | AGFI | 0.909 | The required level is achieved |
| 2. Incremental Fit | CFI | 0.957 | The required level is achieved |
|  | TLI | 0.950 | The required level is achieved |
|  | NFI | 0.932 | The required level is achieved |
| 3. Parsimonious Fit | Chisq/df | 2.588 | The required level is achieved |

RMSEA: Root Mean Square of Error Approximation
GFI: Goodness of Fit Index
AGFI: Adjusted Goodness of Fit
CFI: Comparative Fit Index
TLI: Tucker-Lewis Index
NFI: Normed Fit Index
Chisq/df: Chi Square/Degrees of Freedom

Based on Table 8, the results of the modification model also show that the CR values are above 0.6 and AVE values exceed 0.5, indicating good convergent validity and composite reliability. MSV and ASV values that are smaller than AVE indicate good discriminant validity of the construct. The results shown in Fig 5 illustrate that the final modification model consists of 24 items that retained all six dimensions of the original measurement model by Ajis et al. [21]. For the self-confidence dimension, the model retained only four out of eight items. For the family and close relationships adaptation dimension, the model retained only five out of eight items. For the living environment adaptation and economic stability dimensions, the model retained only four out of five items. The model maintained all three items for the physical health dimension and four items for the autonomy and agency dimension.

**Table 8.**
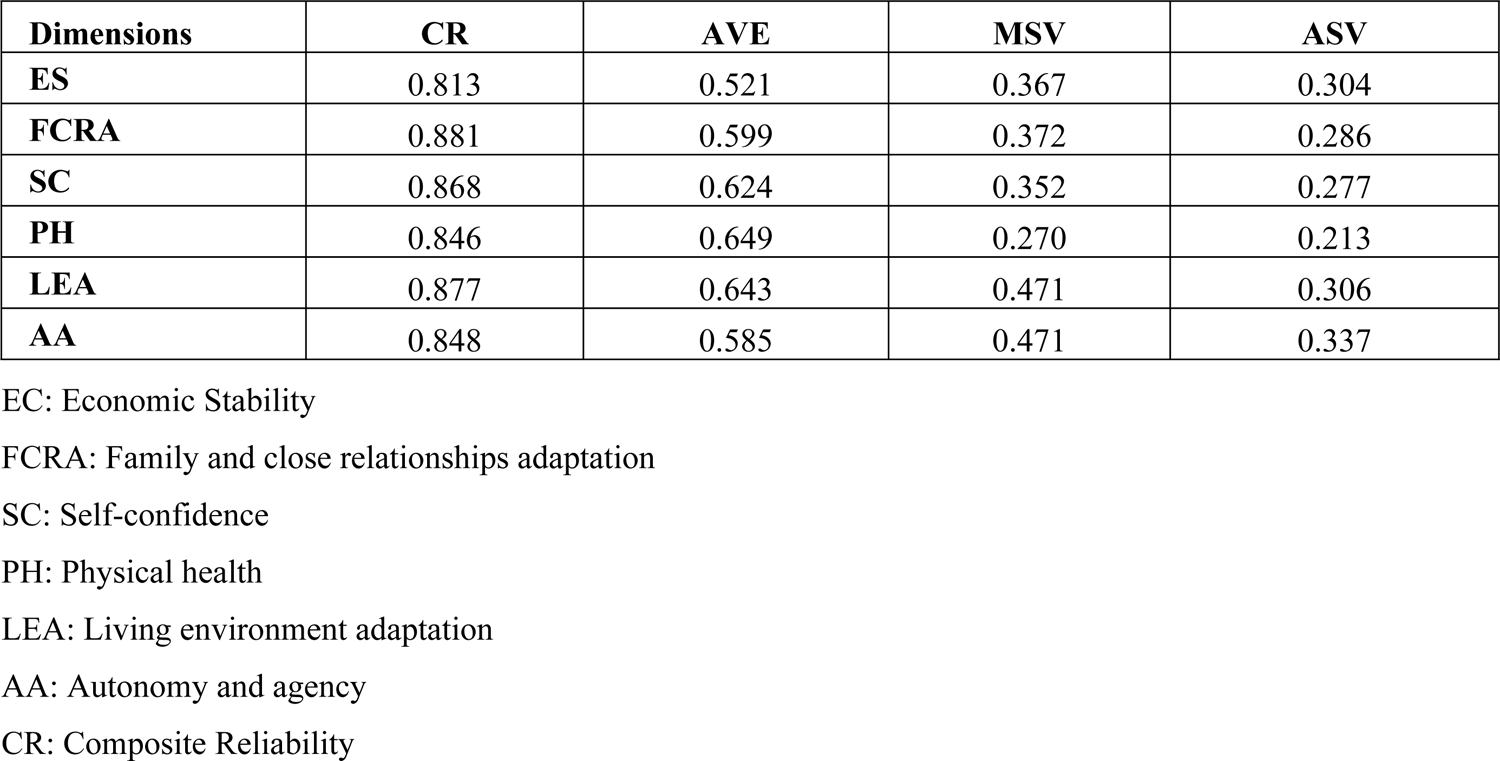
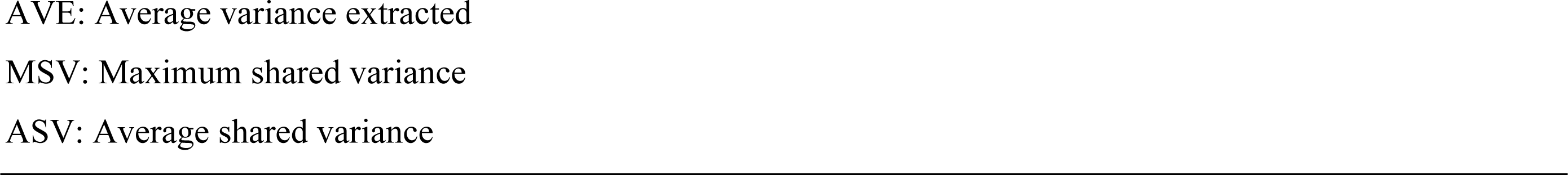
Composite reliability, convergent validity and discriminant validity among dimensions in the final modification model for male well-being.

| Dimensions | CR | AVE | MSV | ASV |
| --- | --- | --- | --- | --- |
| ES | 0.813 | 0.521 | 0.367 | 0.304 |
| FCRA | 0.881 | 0.599 | 0.372 | 0.286 |
| SC | 0.868 | 0.624 | 0.352 | 0.277 |
| PH | 0.846 | 0.649 | 0.270 | 0.213 |
| LEA | 0.877 | 0.643 | 0.471 | 0.306 |
| AA | 0.848 | 0.585 | 0.471 | 0.337 |

EC: Economic Stability
FCRA: Family and close relationships adaptation
SC: Self-confidence
PH: Physical health
LEA: Living environment adaptation
AA: Autonomy and agency
CR: Composite Reliability
AVE: Average variance extracted MSV: Maximum shared variance ASV: Average shared variance

Furthermore, the modification model in Fig 5 also presents the measure of correlation among six dimensions measuring male well-being, illustrating that none of the correlation values between any two dimensions exceeded 0.85. Thus, the modification model does not have multicollinearity problems.

The correlation among dimensions obtained from the modification model in Fig 5 is tabulated in Table 9. It is concluded that the discriminant validity for constructs in the final modification model has been achieved since all diagonal values are greater than any other values in the rows and columns (26–32).

**Table 9.** Discriminant validity index summary for the final modification model.

|  | ES | FCRA | SC | PH | LEA | AA |
| --- | --- | --- | --- | --- | --- | --- |
| ES | 0.722 |  |  |  |  |  |
| FCRA | 0.538 | 0.774 |  |  |  |  |
| SC | 0.593 | 0.541 | 0.790 |  |  |  |
| PH | 0.441 | 0.438 | 0.490 | 0.805 |  |  |
| LEA | 0.561 | 0.610 | 0.455 | 0.408 | 0.802 |  |
| AA | 0.606 | 0.535 | 0.540 | 0.520 | 0.686 | 0.765 |

ES: Economic stability FCRA: Family and close relationships adaptation SC: Self-confidence PH: Physical health LEA: Living environment adaptation AA: Autonomy and agency

This study concludes that the final modification model that has been constructed and tested using CFA has produced a model with good fit indices and a set of items suitable for measuring male well-being in Malaysia. This finding further illustrates that the 24 items used to measure male well-being were suitable with the data of the study.

### Instrument reliability

Table 10 illustrates the summary for instrument reliability through assessment of Cronbach’s alpha values. All the dimensions for male well-being in the final modification model indicate good reliability levels (more than 0.7) across the six dimensions (self-confidence, family and close relationships adaptation, physical health, living environment adaptation, autonomy and agency and economic stability). The construct validity has also been reviewed by observing the Pearson correlation values of each item against the total scores of the measured variables. The result of this study shows that the correlation value of each item with their overall dimensions is high (0.585 to 0.804).

**Table 10.** Cronbach’s alpha values for dimensions in the final modification model.

| Dimensions | 24 Item in Modification Model | Corrected Item-Total Correlation | Cronbach's Alpha if item deleted | Cronbach's Alpha values |
| --- | --- | --- | --- | --- |
| Self-confidence | C1.1 | 0.642 | 0.857 | 0.865 |
|  | C1.2 | 0.745 | 0.814 |  |
|  | C1.3 | 0.774 | 0.803 |  |
|  | C1.4 | 0.699 | 0.833 |  |
| Family and close relationships adaptation | C2.2 | 0.731 | 0.841 | 0.874 |
|  | C2.3 | 0.635 | 0.871 |  |
|  | C2.4 | 0.697 | 0.848 |  |
|  | C2.6 | 0.734 | 0.839 |  |
|  | C2.7 | 0.752 | 0.837 |  |
| Physical health | C3.1 | 0.715 | 0.768 | 0.839 |
|  | C3.2 | 0.752 | 0.730 |  |
|  | C3.3 | 0.653 | 0.835 |  |
| Living environment adaptation | C4.2 | 0.682 | 0.848 | 0.869 |
|  | C4.3 | 0.804 | 0.799 |  |
|  | C4.4 | 0.763 | 0.816 |  |
|  | C4.5 | 0.654 | 0.866 |  |
| Autonomy and agency | C5.1 | 0.711 | 0.816 | 0.858 |
|  | C5.2 | 0.734 | 0.806 |  |
|  | C5.3 | 0.662 | 0.835 |  |
|  | C5.4 | 0.703 | 0.819 |  |
| Economic stability | C6.1 | 0.643 | 0.752 | 0.809 |
|  | C6.2 | 0.662 | 0.742 |  |
|  | C6.4 | 0.585 | 0.78 |  |
|  | C6.5 | 0.631 | 0.763 |  |
| Modification Model |  |  |  | 0.841 |

The final modification model for measuring male well-being was reliable, with high internal consistencies. All sub-scales achieved Cronbach’s alpha of 0.80 and above, and the overall instrument achieved 0.841. These results represent equivalence and consistency among the responses to items of the final modification model, suggesting that these items are suitable for measuring male well-being compared to the original measurement model in the Malaysian context. The internal consistency reliability was robust, with no floor/ceiling effects.

The original measurement model was developed via an exploratory factor analysis (EFA) procedure using pilot study data. Ajis et al. [21] suggested that the instrument should be further validated with additional data from the field. In the present study, the researchers utilised CFA as a procedure for validating this instrument as per common practice [28,32,34–38].

## Conclusion

The modification model developed through this study has resulted in 24 items measuring male well-being in the Malaysian context. This instrument may be used in measuring male well-being in Malaysia as it achieved robust reliability and structural validity that fulfilled goodness-of-fit criteria. It is also deemed suitable to measure male well-being in similar cultural contexts.

This study is a new finding in furthering research related to well-being in the context of gender and cultural norms. It is the first study conducted to develop an instrument for the measurement of male well-being in Malaysia. Dimensions of well-being developed specifically for men can be used to measure and potentially unravel problems involving men’s well-being. Self-report assessments used as a tool in measuring men’s well-being provide insight into the importance of social perspectives that have an impact on men’s well-being. Furthermore, this study adds new knowledge about the construct and conceptualization of men’s well-being in Malaysia. The instrument may be used by individuals, organizations, and government agencies, specifically those with a focus on understanding well-being from the male perspective.

Even so, this study has several limitations. First, the study was conducted through a convenience and snowball sampling strategy using network chains from the research team and spread through different social media platforms (Whatsapp, Facebook, Twitter, and Instagram). As a result, there is a possibility of bias because disadvantaged populations or those with limited access to the internet may not have been able to participate in this study. In addition, when compared to the current statistical population in Malaysia, the study sample overrepresents Malay males. Thus, the findings of this study must be interpreted with caution. A more systematic and inclusive sampling method is needed to improve findings in terms of population representation and generalization.

Secondly, the original study [21] formed indicators of male well-being based on elements of self-concept theory, which may have limited the scope of its definition of the well-being concept. Triangulation of the findings through qualitative interviews would allow respondents to provide more specific details and potential areas of improvement to the instrument. This would be a useful addition to understanding the subjective evaluation of well-being among men for future studies.

To improve the generalizability of findings, more validation studies should be conducted considering this study’s limitations. The model should also be tested in different cultural settings to observe its validity and reliability in different contexts.

## Data Availability

All data files are available from the Harvard Dataverse database (doi:10.7910/DVN/ESH83R).

https://dataverse.harvard.edu

## Acknowledgements

This study was funded by research grant, Center for Research and Instrument Management, Universiti Kebangsaan Malaysia (GUP-2016-064) and also the Ministry of Women, Family and Community Development Malaysia (SK-2016-002). We also like to express our thanks to Assoc. Prof. Dr. Mohammad Rezal Hamzah and Mr. Andi Muhammad Tri Sakti for their assistance throughout this study.

## Supporting information

### Author Contributions

Conceptualization: EM, AAA and SZMA

Formal Analysis: SZMA

Supervision: EM and AAA

Writing - Original Draft Preparation: SZMA

Writing - Review & Editing: AAA and EM

